# Exploring Vaccine Hesitancy in Care Home Employees in North West England: A Qualitative Study

**DOI:** 10.1101/2021.08.20.21262101

**Authors:** Amelia Dennis, Charlotte Robin, Leah Jones, Holly Carter

## Abstract

**Objectives:** Care homes have experienced a high number of COVID-19 outbreaks, and it is therefore important for care home employees to receive the COVID-19 vaccine. However, there is vaccine hesitancy at a greater rate among this group than in the wider community. We aimed to understand barriers and facilitators to getting the COVID-19 vaccine, as well as views on potential mandatory vaccination policies.

**Design:** Semi-structured interviews.

**Setting:** Care home employees in North West England. Interviews conducted in April 2021.

**Participants:** 10 care home employees (aged 25 to 61 years old) in the North West, who had been invited to have, but had not yet received, the COVID-19 vaccine.

**Results:** We analysed the interviews using a framework analysis. Our analysis identified eight themes: perceived risk of COVID-19, efficacy of the vaccine, concerns about the vaccine, mistrust in authorities, facilitators to getting the vaccine, views on potential mandatory COVID-19 vaccination policies, negative experiences of care work during the COVID-19 pandemic, and communication challenges.

**Conclusions:** The care home employees interviewed at the time of this study reported a low perceived risk of COVID-19, alongside concerns over the efficacy of the vaccine, side effects, and speed of vaccine development, which is exacerbated by mistrust in authorities. Facilitators to getting the vaccine included wanting to protect others, perceived severity of COVID-19, and workplace norms. Making COVID-19 vaccination a condition of deployment may not result in increased willingness to get the COVID-19 vaccination, with most care home employees in this study favouring leaving their job rather than getting vaccinated. At a time when many of the workers already had negative experiences of care work during the pandemic due to perceived negative judgment from others and a perceived lack of support facing care home employees, policies that require vaccination as a condition of deployment were not positively received.

## INTRODUCTION

The COVID-19 pandemic, caused by the spread of SARS-CoV-2, has led to substantial mortality and strain on health and care systems. One population most at risk of COVID-19 is care home residents and employees. Care homes have been disproportionately affected by COVID-19, with high rates of illness and death among care home residents, particularly in the first wave of the pandemic. Investigations into local outbreaks revealed that in April 2020, 21% of care home workers tested positive for COVID-19 [1], and 31.1% of deaths in the first wave occurred in care home residents [2]. In some regions over a third of care homes have experienced an outbreak [3].

In 2020, several vaccines were developed that were shown to be effective and safe [4], reducing both hospitalisation and mortality [5]. Consequently, several COVID-19 vaccines were authorised for use in multiple countries including the UK [6]. It has been estimated between 55% to 85% of populations would need to be vaccinated in order to reach herd immunity for COVID-19, depending on infection rates within each country [7, 8].

When England commenced COVID-19 vaccination rollout on the 8^th^ December 2020, care home residents and employees were in the first priority group to receive the vaccine. By 15^th^ February 2021, all care home residents and employees had been offered their first dose of COVID-19 vaccine. However, some care homes saw low COVID-19 vaccine uptake among employees. As of the 29^th^ April 2021, 94.8% of eligible care home residents and 81.0% of eligible care home employees had received their first COVID-19 vaccination [9]. Vaccine uptake among care home employees differs regionally, with the North East and South West having vaccine uptake above 84%, and the North West and London having vaccine uptake below 79%. In some areas in the North West reported vaccine uptake amongst care home employees is much lower, for example, a recent survey of care home staff in Liverpool identified vaccination uptake was at 51.4% [10]. Therefore, it is important to understand reasons behind the lower vaccination uptake among some care home employees.

Research has identified several barriers to getting the COVID-19 vaccine, including mistrust in healthcare professionals and scientists, safety concerns, negative perceptions of vaccination side effects, lower perceived threat of COVID-19, and inconsistent information [11-15]. Barriers for care home employees include perceived lack of research, concerns over fertility, concerns over allergic reactions, and difficulty accessing vaccination appointments [10], with similar barriers also being identified among healthcare workers [16]. Despite these barriers, a recent meta-analysis of 40 studies found 61% of all healthcare workers agreed with mandatory vaccines, although vaccinated healthcare workers were more likely to favour mandatory vaccine than unvaccinated healthcare workers [17].

### The current study

Care home employees are at a higher risk of contracting COVID-19 but are also a population with lower COVID-19 vaccination uptake. On 14^th^ June 2021, the UK government announced it would be a condition of deployment for care home employees working in older adult care homes in England to receive COVID-19 vaccination, and they would have 16 weeks to do this [18]. In the current study, we explored barriers to COVID-19 vaccination uptake among care home employees, including perceptions of mandatory COVID-19 vaccination (prior to the announcement COVID-19 vaccine would become a condition of deployment for care home employees). We used a qualitative method that allows for in-depth exploration of personal experience [19] and provides rich detail on feelings and experiences behind attitudes [20].

The current study had three aims. First, to identify reasons underlying care home employee decisions to decline COVID-19 vaccination. Second, to explore any factors that might increase vaccine uptake among care home employees. Third, to identify attitudes towards mandatory vaccinations.

## METHOD

### Participants

Ten participants took part, aged 25-61 years old, see Table 1 for demographics. Participants were invited to take part in the study if they worked in a care home and had decided not to get their COVID-19 vaccination. Participants were recruited by contacting Health Protection leads for the North West, who then used snowballing sampling to identify local authorities who would be willing to assist with recruitment. The lead researcher then contacted the engaged local authorities and the four that agreed to help contacted care homes in their region and passed on the recruitment information. Participants then opted-in to the study by contacting the lead researcher using the details provided. All participants received a £20 voucher for taking part.

**Table 1.** Participant demographics.

|  | Frequency | Percentage |
| --- | --- | --- |
| Age | $M=38.20$ | $SD=12.23$ |
| Gender |  |  |
| Female | 7 | 70% |
| Male | 3 | 30% |
| Employed by |  |  |
| Agency | 1 | 10% |
| Care Home | 9 | 90% |
| Type of Care Home |  |  |
| Disabilities | 1 | 10% |
| Home Carer | 2 | 20% |
| Mental Illness | 3 | 30% |
| Residential | 4 | 40% |

### Patient and public involvement

There was no patient or public involvement.

### Interview schedule

We developed the interview schedule through discussions based on prior knowledge and existing research [21]. The interview schedule centred around three main topics. First, barriers and facilitators to getting COVID-19 vaccination when offered. Second, sources of information about COVID-19 vaccination. Third, views on mandatory vaccinations. Open-ended questions and prompts guided the interviews to develop a conversational style to elicit rich descriptions [22].

### Procedure

Potential participants contacted the researcher and were given a copy of the information sheet. In some cases, details of potential participants were provided by care homes or local authorities and the information sheet was sent to them directly. Participants were sent an online survey to provide informed consent and basic demographic information. The first author conducted all interviews, which took place online during April 2021, before the announcement of potential mandatory vaccination for care home employees. Interviews ranged between 26 and 65 minutes and each interview was recorded and later transcribed.

### Data analysis

We analysed the data using framework analysis as it is grounded in data, flexible, and has been used widely to inform policy [23]. Using NVivo, the five steps of framework analysis were applied to the data [24]. In step one, interview transcripts were read; this familiarisation was then used to inform step two whereby the researcher identified codes in the data that related to the research questions, which included barriers to getting COVID-19 vaccine. Indexing was then carried out, whereby data were identified that related to broad themes which were then discussed with other members of the research group. Step four involved charting the data by summarising it into an analytic framework. All transcripts were coded by the lead researcher, with 30% double-coded by a second researcher to ensure consistency. The team then met to discuss the charted data, which led to the final themes. In the final step, themes were defined and clarified in relation to other themes. On completion of the analysis, no new themes emerged from the data and data saturation had been reached [25, 26].

## RESULTS

Eight main themes were identified: perceived risk of COVID-19, efficacy of the vaccine, concerns about the vaccine, mistrust in authorities, facilitators to getting the vaccine, views on mandatory vaccinations, negative experiences of care work during the COVID-19 pandemic, and communication challenges. An overview of the themes and sub-themes is presented in Table 2.

**Table 2.**
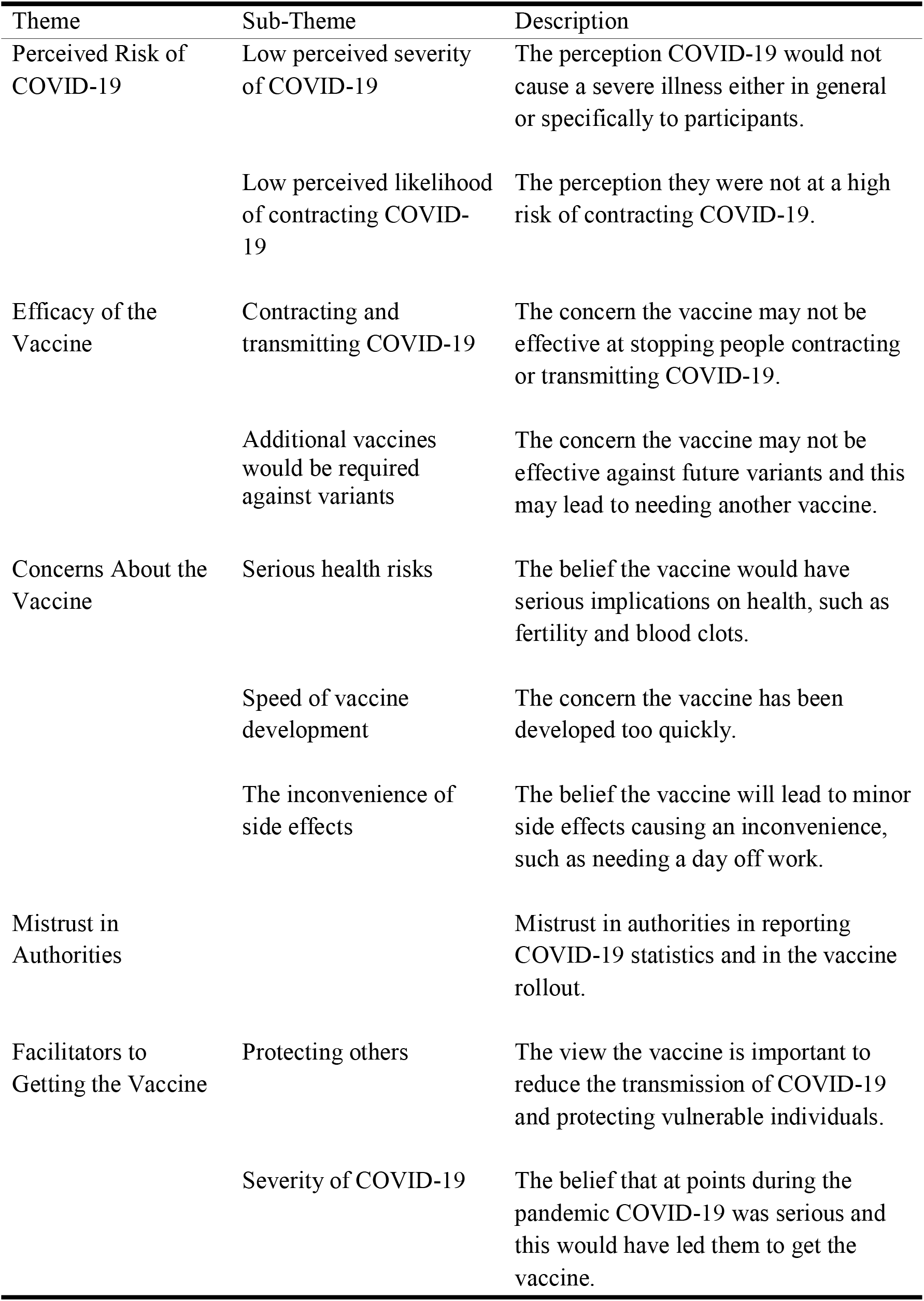
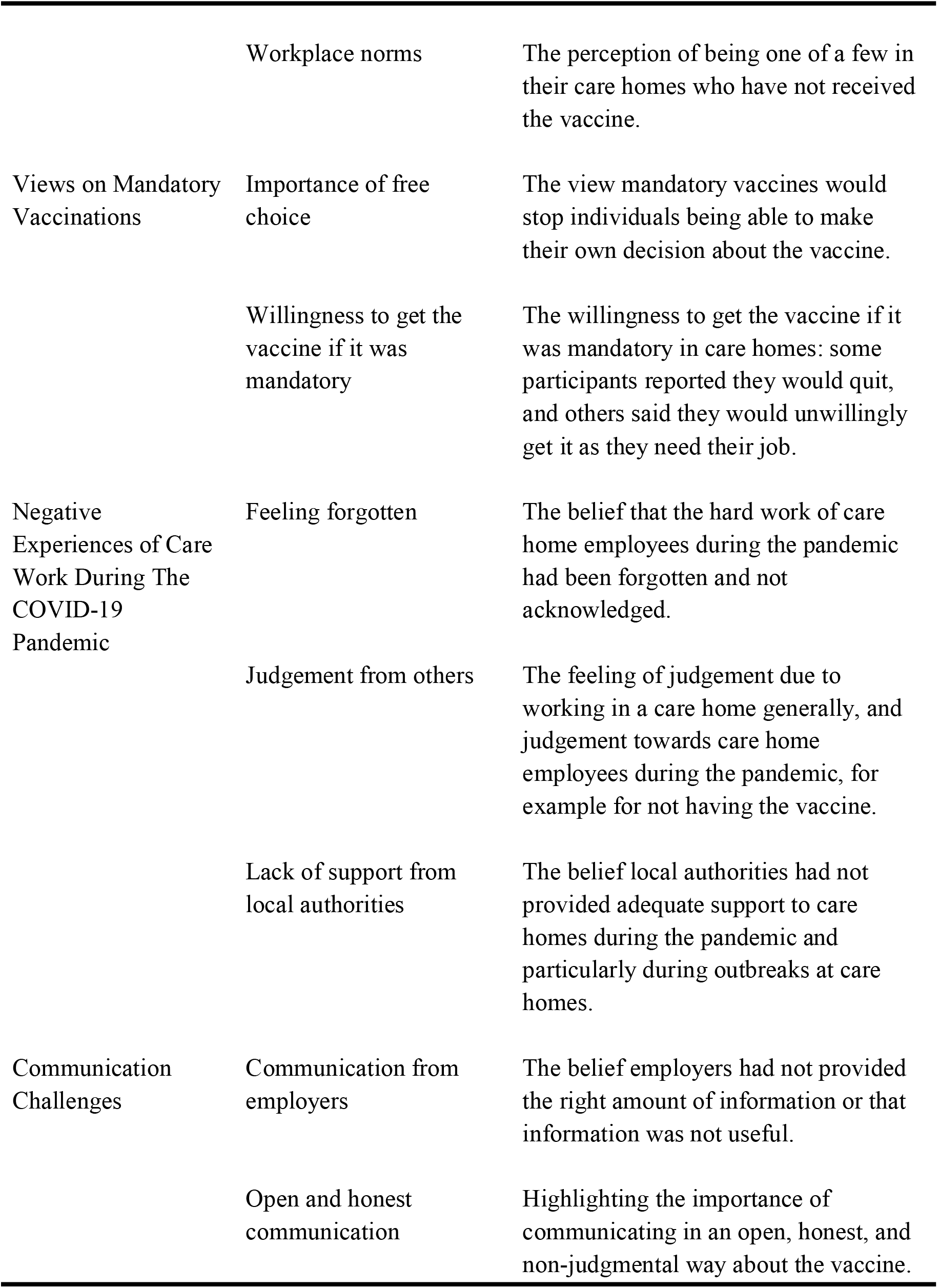
Overview of the themes and sub-themes.

| Theme | Sub-Theme | Description |
| --- | --- | --- |
| Perceived Risk of COVID-19 | Low perceived severity of COVID-19 | The perception COVID-19 would not cause a severe illness either in general or specifically to participants. |
|  | Low perceived likelihood of contracting COVID-19 | The perception they were not at a high risk of contracting COVID-19. |
| Efficacy of the Vaccine | Contracting and transmitting COVID-19 | The concern the vaccine may not be effective at stopping people contracting or transmitting COVID-19. |
|  | Additional vaccines would be required against variants | The concern the vaccine may not be effective against future variants and this may lead to needing another vaccine. |
| Concerns About the Vaccine | Serious health risks | The belief the vaccine would have serious implications on health, such as fertility and blood clots. |
|  | Speed of vaccine development | The concern the vaccine has been developed too quickly. |
|  | The inconvenience of side effects | The belief the vaccine will lead to minor side effects causing an inconvenience, such as needing a day off work. |
| Mistrust in Authorities |  | Mistrust in authorities in reporting COVID-19 statistics and in the vaccine rollout. |
| Facilitators to Getting the Vaccine | Protecting others | The view the vaccine is important to reduce the transmission of COVID-19 and protecting vulnerable individuals. |
|  | Severity of COVID-19 | The belief that at points during the pandemic COVID-19 was serious and this would have led them to get the vaccine. |
|  | Workplace norms | The perception of being one of a few in their care homes who have not received the vaccine. |
| Views on Mandatory Vaccinations | Importance of free choice | The view mandatory vaccines would stop individuals being able to make their own decision about the vaccine. |
|  | Willingness to get the vaccine if it was mandatory | The willingness to get the vaccine if it was mandatory in care homes: some participants reported they would quit, and others said they would unwillingly get it as they need their job. |
| Negative Experiences of Care Work During The COVID-19 Pandemic | Feeling forgotten | The belief that the hard work of care home employees during the pandemic had been forgotten and not acknowledged. |
|  | Judgement from others | The feeling of judgement due to working in a care home generally, and judgement towards care home employees during the pandemic, for example for not having the vaccine. |
|  | Lack of support from local authorities | The belief local authorities had not provided adequate support to care homes during the pandemic and particularly during outbreaks at care homes. |
| Communication Challenges | Communication from employers | The belief employers had not provided the right amount of information or that information was not useful. |
|  | Open and honest communication | Highlighting the importance of communicating in an open, honest, and non-judgmental way about the vaccine. |

### Perceived risk of COVID-19

Participants views on the risk of COVID-19 were centred around the severity of COVID-19 and the likelihood of contracting COVID-19.

#### Low perceived severity of COVID-19

Many participants perceived the severity of COVID-19 to be low and thought the risk had been exaggerated: “*For me it’s just blown out of proportion*” (Participant 1). Other participants felt that while COVID-19 could be severe for some, it would not be severe for them: “*I’m probably a lot less likely to get quite ill if I got coronavirus*” (Participant 7). This belief was often due to positive perceptions of their own health: “*I just thought the chances of me actually getting ill are very slim anyway, I’m quite healthy in myself anyway, I’m active*” (Participant 10), or to their mindset in dealing with pain and illness: “*I’m a pretty strong person. I can take pain. I’m never sick. I don’t do sick and stuff like that. It seems to be a mind-set as well*” (Participant 3). Additionally, participants perceived COVID-19 as not severe as they already had antibodies: “*I know that I already have antibodies to COVID and after doing some reading on it, it seems as though the antibodies do last a long time so hopefully that will provide me with some immunity*” (Participant 4), or because of natural immunity: “*I don’t take any vaccinations. I believe that we have an immune system*” (Participant 6).

The view severity of COVID-19 was low reduced intentions towards getting the vaccine, as their perceived risk of the vaccine outweighed their perceived risk of COVID-19: “*I know the pros of having the vaccine outweigh the risk, but are you willing to take that risk when the chances of even getting COVID and having a bad reaction to it are low anyway*” (Participant 6). However, it was acknowledged that if they had underlying health conditions, they would be more likely to get the vaccine: “*if I had any illness or ongoing issues such as asthma, COPD, or like I was at a certain age category, I would have taken it*” (Participant 10).

#### Low perceived likelihood of contracting COVID-19

The participants who had not contracted COVID-19 perceived their risk of catching it as low. Some attributed this to the fact they had not yet caught COVID-19: “*if I was gonna get COVID I would have got it*” (Participant 3), while others felt the precautions they had taken would reduce their likelihood: “*I take enough precautions. Washing hands, keeping distance, and keeping mask on all the time”* (Participant 3).

### Efficacy of the vaccine

Participants’ concerns about the efficacy of the vaccine centred around two different sub-themes: efficacy of the vaccine for preventing people from contracting and transmitting COVID-19, and the anticipation additional vaccines would be required to protect against variants.

#### Contracting and transmitting COVID-19

Several participants felt the COVID-19 vaccine would not be effective. The reasons given for this were a belief the COVID-19 vaccine does not lower the chances of transmitting COVID-19: “*it doesn’t stop from spreading it anyway*” (Participant 2), and that more time is needed to understand how effective the vaccine will be: “*at the minute we’re not even 100% sure the vaccine is gonna work […]We’ll only really find out end of this year, if it’s worked or not”* (Participant 10). Views about vaccine efficacy were drawn from two main sources. The first was secondhand experiences: “*I know people who have taken the vaccine and still got positive* … *so it’s not a cure*” (Participant 3), and the second was information they had read: “*It’s not been proven that if you have the vaccine you can’t pass it on, they’ve said you can still catch it and you can still pass it on*” (Participant 4).

#### Additional vaccines would be required against variants

Another concern for participants was the different variants of the COVID-19 virus. Participants suggested a new vaccine may have to be developed to protect against variants, and this made them reluctant to have COVID-19 vaccination: “*with the new variants coming out they are still tweaking it*” (Participant 3). This resulted in participants believing that even if they got the COVID-19 vaccination now, they may still have to get more vaccinations in the future: “*This vaccine it’s meant to be COVID-19 right but everyday there is a new variant … does that mean we are taking all of these vaccines to keep up with it*” (Participant 1). Another participant questioned the efficacy of the different types of the COVID-19 vaccination in relation to the different variants: “*the different variants […] we say this vaccine doesn’t work against that one, that one doesn’t work against that one*” (Participant 10).

### Concerns about the vaccine

All participants mentioned concerns about the COVID-19 vaccine, which influenced their decision not to be vaccinated. These concerns could be broadly grouped into three sub-themes: serious health risks associated with the vaccine, the speed of vaccine development, and the inconvenience of side effects.

#### Serious health risks

One of the key barriers was concern about the serious health risks the vaccine may cause: “*it’s just the health risks that is a concern for me*” (Participant 9). One participant mentioned their allergy causing them to be at greater risk of experiencing adverse effects: “*I am anaphylactic and I know originally the Pfizer one was deemed anyone anaphylactic […] to postpone getting the vaccine […] that’s another no from me*” (Participant 6). This theme links with perceived severity of COVID-19, participants mentioned that for them, health risks posed by the vaccine outweighed any risk of contracting COVID-19: “*the risks for me with having the vaccine outweigh the risks I’d actually get if I got COVID myself”* (Participant 10).

Participants’ beliefs about health risks associated with the COVID-19 vaccine were drawn from two main sources. First, the health risks of the vaccine were influenced through family experiences of getting the vaccine: “*My mum had the vaccine she was more unwell from that than COVID. She was in hospital, she was really unwell, but she was not that unwell from COVID*” (Participant 9). Second, health risks of the COVID-19 vaccine have been widely reported in the media, particularly in regards to blood clots: “*there were things coming up about blood clots and stuff and it was kind of another push in the direction of not really wanting it*” (Participant 7), and concerns about impact on fertility: “*at the time I had heard mentions about pregnancy with having the vaccine, so I spoke to my GP and they kind of I think erred on the side of caution saying urm maybe not”* (Participant 7).

#### Speed of vaccine development

A lot of participants expressed concern about the speed with which the COVID-19 vaccine has been developed and rolled out: “*I’m just not comfortable with the fact it’s only just been developed*” (Participant 5). The speed of development had led to mistrust in the vaccine rollout, specifically in the testing surrounding the vaccine.

> “*I do feel uncomfortable about how quickly it’s been rolled out […]they say when they roll it out that it has been through all the tests and stages that it would usually go through […] But there’s been a few things since then that have just made me question whether that really is accurate*.” (Participant 4)

In addition, some participants expressed hesitancy about getting vaccinated because they wanted to wait for further testing of the vaccine to be done. This linked with wanting the vaccine in future, when further testing had been completed.

> “*I want to personally wait […] until the testing had been fully done and then I was gonna make a decision you know just by being cautious*.” (Participant 10)

Some participants specifically stated that the speed of development of the COVID-19 vaccine had exacerbated their concerns about the safety of the vaccine: “*just the way it was rushed and then you’ve got people dying of blood clots*.” (Participant 1)

#### The inconvenience of side effects

Alongside concerns about serious health risks, another concern was the inconvenient side effects of the COVID-19 vaccine that did not pose a serious threat to health, but rather were unpleasant or inconvenient. Some participants mentioned side effects their family or friends experienced after receiving the vaccine: “*I can’t think of anyone who hasn’t had side effects from it*” (Participants 4). Additionally, one participant explained they were concerned about the side effects as they did not want to get ill and take time off work: “*I won’t take time off work*” (Participant 3).

### Mistrust in authorities

Participants expressed a lack of trust in authorities that contributed to their hesitancy in getting the COVID-19 vaccine. There were several reasons for mistrust in authorities in relation to COVID-19. First, there was scepticism about the way COVID-19 deaths were reported: “*people who are testing positive for COVID even if they are dying by a bus hitting them, they are being marked down as COVID related”* (Participant 1). Second, there was mistrust over hospitalisation statistics: “*But when they’re saying the COVID beds are at full capacity, I know people that work in hospitals […] The hospital’s not overrun*” (Participant 6). Third, there was mistrust due to the perceived poor support provided to care homes: “*there was little to no help from the government. And in terms of PPE I think we were completely and utterly failed in terms of PPE*” (Participant 8).

Mistrust in authorities has contributed to safety concerns about the vaccine: “*The NHS have done some dodgy things over the years and the government, the blood scandal, the thalidomide scandal, and it’s just like this is not tested*” (Participant 5); “*only given emergency approval so it’s not even got the full 100% safety approval by any European standards*” (Participant 6). Participants also specifically highlighted lack of trust as a reason for not getting the vaccine: “*Normally vaccine takes time to sort out. But this is done quickly, cut corners. You know you can’t trust anybody these days, you know scientists, so I thought I’ll wait*” (Participant 3).

### Facilitators to getting the vaccine

Although participants did not get the vaccine, they did mention reasons to get the vaccine. These facilitators were wanting to protect others, the severity of COVID-19, and workplace norms.

#### Protecting others

The main reason participants thought they should get the vaccine was to protect others and reduce COVID-19 transmission.

> Interviewer: “*So if you did get the vaccine, would it be for yourself or would it be to stop you transmitting Covid to others?*”
>
> Participant 10: “*Stop transmitting to others, it wouldn’t be for myself*.*”*

There was an understanding of how the vaccine can reduce COVID-19 transmission due to the reduction in cases since the start of the vaccine rollout: “*It protects people against the virus. Because it’s not just to protect you. It’s to protect other people. It seems like it is the right thing to do because the rates are going down since people have been vaccinated*.” (Participant 9).

However, others highlighted the need for vulnerable people to protect themselves with the vaccine: “*So, if all the residents have been vaccinated, which they now have, I don’t see why it would make any difference as to why the carers would need it*” (Participant 4).

#### Severity of COVID-19

The final reason why participants thought they should get the vaccine was because they perceived COVID-19 to be serious, which was particularly relevant at the beginning of the pandemic.

> Interviewer: “*Are there any reasons you’ve thought you should get the vaccine?”* Participant 1: “*Right at the beginning when we started hearing about COVID, and it was killing people, people were dying from COVID, that was a concern because it was like, oh, hang on a second*.*”*

When participants perceived COVID-19 to be severe they would have been more likely to accept the COVID-19 vaccine to prevent severe consequences.

> “*If there was a vaccine right at the beginning, I probably would have taken it. Only because it was people are dying. People are dying, and this is happening. We have, not a cure, something which will stop you ending up being on ventilator*.” (Participant 1)

#### Workplace norms

There was a workplace norm of getting the COVID-19 vaccine and participants who did not get it were in the minority: “*the majority of people I work with have had it, yes*” (Participant 7). In some instances, this workplace norm did influence them to book their appointment but did not lead to them getting the vaccine.

> “*Everyone were talking about having it and stuff like that and there were saying do you want to put your name on? And obviously everyone is round the table because I’m on our break and I’m like yes okay*” (Participant 10).

### Views on mandatory vaccinations

Participants’ views about mandatory vaccination could be grouped into two main sub-themes: the importance of free choice; and willingness to take a mandatory vaccination for work purposes.

#### Importance of free choice

All participants felt it was important to have the freedom to decide whether to have the vaccine or not: “*It’s the wording of, it’s the way that it’s been pushed upon people which I disagree*” (Participant 1). The narrative of mandatory vaccines negatively influenced participants’ attitudes towards the vaccine: “*now that the government want to try and make us have it that’s made me feel really like I don’t want to have* … *you can’t force people to do what they don’t want to do*” (Participant 5).

There were several reasons why participants felt mandatory vaccination was inappropriate. First, making vaccines mandatory would cause a perceived lack of transparency: “*people can make the decision for themselves on the benefits to risks ratio and people weren’t given the opportunity to do that, so I think there’s been a real lack of transparency and that’s a concern*” (Participant 5). Second, because it would prevent those who had not been vaccinated from certain freedoms: “*I feel that it’s terrible that you might not be able to do things because you’ve not had the vaccine*” (Participant 9). Last, that it would be a betrayal of care home employees who have worked in care for years.

> “*Most staff in care they work usually for over 10 years in care so I think it’d be a bit disheartening to put that much time into it and then they just get dismissed because they’ve not had a vaccine especially when there’s not loads of evidence yet I think it’d be a bit cruel*.” (Participant 10)

#### Willingness to get the vaccine if it was mandatory

All participants expressed their anger towards the possibility of vaccines being mandatory among care home employees: “*I’d be absolutely livid … I think that would be disgusting”* (Participant 6). There were a range of responses in terms of what participants would do if vaccines were made mandatory for care home employees. Some said they would leave their job so they did not have to get the vaccine: “*well if it was mandatory for people who are already employed in that area then I’d be out of a job*” (Participant 2). This was true even for some who expressed how much they enjoyed their jobs.

> “*Since they’ve been talking about [mandatory vaccinations] Since that moment*… *I love my job in the care home and I’d planned to stay there a long time, but I have been looking for a new job*.*” (Participant 4)*

Others said they would reluctantly have the vaccine so they can keep their job: “*it’s either that or lose my job and not be able to pay my rent and buy food for my kids*” (Participant 5). One participant said they would try to avoid having it via a medical exemption: “*I am anaphylactic so [that’s] how I would get out*” (Participant 6).

### Negative experiences of care work during the COVID-19 pandemic

Participants generally reported negative experiences of working in a care home during the pandemic, with sub-themes including: feeling forgotten; experiencing judgement from others; and a lack of support from local authorities.

#### Feeling forgotten

There was a strong sense of feeling forgotten and underappreciation of care home employees during the pandemic: “*Like I thought we might have been given something because we’ve had to work through the pandemic, but we literally received nothing in that way*” (Participant 10). Participants felt that while the efforts of NHS staff had been recognised, they had been forgotten.

> “*Nobody’s acknowledged what the care homes actually had to go through, and how hard we’ve all worked as well. It’s just the NHS that gets prioritised, when they weren’t even treating these elderly people most of the time*.” (Participant 5) “*We got no help. I’m not taking anything away from what the NHS staff did, they did amazing work, but they were getting all the recognition*.” (Participant 9)

#### Judgment from others

However, many participants reported not just feeling forgotten, but experiencing negative judgement from the general public. This included negative judgements about care work in general: “*People like carers on, like I said, one of the lowest paid jobs in the country. I think we’re so disposable to them*” (Participant 9); “*when you’ve got people just criticising you, putting you down in your job when you’re just trying to look after people, it’s quite demoralising*” (Participant 10).

There was also perceived judgement about aspects of the COVID-19 response, such as care home outbreaks: “*At the beginning there was the blame that the carers are bringing it in*” (Participant 8), not letting visitors in: “*Most care homes have COVID. And even now we’re getting slated because we don’t want to let visitors back in, because we’re concerned, and it’s inhumane*” (Participant 5), and not getting the COVID-19 vaccine.

> “*To see them turn on us now and say […] if you’re not going to have the vaccine and you shouldn’t be allowed to go into work until the pandemic is over is quite shocking really and very disappointing*.” (Participant 4)

#### Lack of support from local authorities

Participants described how they felt they had been let down by local authorities during the COVID-19 pandemic. The lack of support by local authorities was highlighted during COVID-19 outbreaks at care homes: “*it (COVID-19) got in and once it got in we had no help from our local council*” (Participant 8). It was believed that the support could have been better coordinated in order to save time during a crisis: “*four different departments were ringing from the council and I was like why are you not liaising with each other, why are there four different people ringing us when we are in crisis*” (Participant 5).

As a result of these negative experiences, some participants expressed a desire to leave their profession: “*Oh yes. I’m currently looking for a new job*” (Participant 9). It was also mentioned that other employees in their care home were looking for new jobs as well: “*Quite a lot of the staff at the home that I currently work at are considering leaving, it’s not public knowledge, but in various different homes a lot of staff are thinking of leaving*” (Participant 8).

### Communication challenges

Some participants discussed areas for improvement in communication, including communication from their employer, and ability to communicate openly with colleagues.

#### Communication from employers

Participants expressed mixed opinions on whether they needed more information from their employers. Some participants wanted more information about the vaccine, particularly in relation to safety concerns.

> “*I think definitely there needs to be more information out there for women who are thinking of fertility is their fertility at risk if they do have the vaccine*.” (Participant 8) *“if the vaccine is causing the blood clots that’s my main concern […] they said it wasn’t related and that it wasn’t to do with it and then said that it was but the chances were very small and then in the next sense they are saying that we are no longer offering it to people under 30*.” (Participant 9)

Other participants felt they had received too much information: “*too much information, it depends where the information is coming from, at the moment I think there is enough information out there*” (Participant 3), and that more information would be unlikely to encourage them to get the vaccine: “*I think more information isn’t going to change my mind at the moment*” (Participant 3). Additionally, some believed the information they had received from their employer was one-sided and needed to be more honest: **“***have to show people the good and the bad but it was a very one sided view point, I don’t think she put any negatives in there*” (Participant 6).

#### Open and honest communication

Some participants felt unable to talk honestly with colleagues about their views on getting the vaccine.

> “*most of the conversation is all of them saying they’ve had theirs and how important it is. I don’t want to negate that, but I don’t also want to get into an argument about why I haven’t*.” (Participant 7)

Others were banned from discussing the vaccine at work for fear of persuading others not to get it.

> “*[My manager] doesn’t want me to discuss it with other staff. Especially when there were five staff at the beginning not wanting it. She didn’t want me to say anything to them, because she really wanted them to have it*.” (Participant 6)

However, others reported that they felt able to discuss their views on the vaccine with colleagues, and this was important for enabling open communication.

> “*Yes everybody has discussions on it, everybody’s got mixed feelings about having it, or mixed reasons for having it. And nobody’s said to anybody else that I can’t believe you’re not having it […] everybody’s quite open to it’s your own decision, if you want it you should have it*.*”* (Participant 8)

## DISCUSSION

In this study, we explored care home employees’ attitudes and behaviours towards the COVID-19 vaccine. Specifically, we examined barriers and facilitators to vaccine uptake and attitudes towards mandatory vaccines. It has since been made a condition of deployment for all staff working in older adult care homes in England to receive both COVID-19 vaccinations [18]. The results from this study therefore provide a timely and important understanding of some attitudes towards and possible consequences of a mandatory vaccination policy, as well as identifying barriers and facilitators to uptake of COVID-19 vaccines.

### Barriers to getting the vaccine

The first aim of this study was to understand barriers to care home employees getting COVID-19 vaccination. Reasons for low vaccine uptake included low perceived risk from COVID-19, low perceived efficacy of the vaccine, concerns about the safety of the vaccine, and mistrust in authorities.

Care home employees perceived low risk from COVID-19, both in terms of the perceived severity of COVID-19 and their perceived likelihood of contracting COVID-19. This led to participants perceiving the risk of having the vaccine (i.e. side effects) as outweighing the benefits (in terms of protecting them from COVID-19). In-line with this, participants stated that if they had an underlying health condition, they would be more likely to get the vaccine due to the increased risk of COVID-19. This is consistent with previous research where individuals with low perceived risk of COVID-19 have increased vaccine hesitancy [14], as they feel less threatened by the disease and thus have less motivation to engage in more preventive behaviour [27].

Care home employees also questioned the efficacy of the vaccine for reducing the risk of transmitting and contracting future variants of COVID-19; as well as the safety of the vaccine, with concerns around the health risks; the speed of development and the inconvenience of side effects. These concerns over the safety of the vaccine were heightened by media reports and the negative experiences of others. Previous research [10, 11] has highlighted concerns over lack of research and a perception vaccine development had been rushed, as reasons for people not getting a COVID-19 vaccine. Participants in the current study reported they would consider getting the vaccine in the future once they perceived it to be effective and safe. This supports previous research, which identifies concerns over lack of research and speed of development as being common reasons for individuals to delay getting their COVID-19 vaccine [16].

Mistrust in authorities contributed to increased concerns about the safety of the vaccine and led to lower perceptions of COVID-19 risk. A lack of trust in the authorities involved in the vaccine response, and in the perceived efficacy and safety of the vaccine, all contribute to a lack of confidence in the vaccine [28]; in turn, lower confidence in the vaccine predicts reduced intentions to get the COVID-19 vaccine [29].

In Liverpool, care home managers have attempted to address COVID-19 vaccine hesitancy through information and myth busting about the vaccine and educational material [10]. However, in this study, care home employees reported not engaging with the information as they felt it was either too much information, not balanced information, or it did not address their specific concerns. Care home employees wanted to discuss the vaccine in an open and honest way where both risk and benefits are acknowledged. Studies have shown providing too much information or unbalanced information can increase vaccine hesitancy [30, 31].

### Facilitators to getting the vaccine

The second aim of the current study was understanding facilitators to care home employees getting a COVID-19 vaccine. Reasons given for potentially getting the vaccine included protecting others, perceived higher severity of COVID-19, and workplace norms. Protecting vulnerable individuals via reduced likelihood of COVID-19 transmission has previously been identified as a facilitator to getting a COVID-19 vaccine among healthcare workers [29]. Some participants highlighted there were times during the pandemic when they perceived COVID-19 to be more serious and as a result would have had the vaccine had it been available. The final facilitator was workplace norms. Most participants reported there was a workplace norm associated with getting the COVID-19 vaccine, and that they were one of few in their workplace who had not received the vaccine.

### Views towards mandatory vaccinations

The final aim of this study was to explore care home employees’ attitudes towards mandatory vaccinations. All participants were strongly opposed to mandatory vaccines, with the media reports in early April 2021 about making the COVID-19 vaccination a condition of deployment for care home workers having a negative impact on participants’ attitudes towards the vaccine. The strong opposition to such policies resulted from a perception that care home workers were not being allowed to make individual health-based decisions based on their own risks. Several participants also believed these types of policies were a betrayal of care home employees who they felt had worked tirelessly during the pandemic with little reward. The lack of acknowledgement and a feeling of being forgotten was frequently raised by participants, who felt they had faced judgement from the public and the media, and a lack of support from local authorities. In response to the possible requirement for care home employees to receive COVID-19 vaccination as a condition of deployment, some participants stated they would leave their job rather than getting the vaccine, whereas others stated they would unwillingly get vaccinated as they need their job. This is in line with previous research that shows the majority of vaccine hesitant individuals do not approve a mandatory policy for the COVID-19 vaccine [32]. Additionally, mandating vaccines can increase anger and negative attitudes among vaccine hesitant individuals, leading to reduced vaccination intentions and uptake [33-36], and potential stigmatisation of those who refuse to have the vaccine [37].

### Recommendations

Based on the findings from this study there are several recommendations that can be made. Firstly, information provided to care home employees should include both the benefits and any risks associated with getting the COVID-19 vaccine. Secondly, employers should facilitate open and non-judgmental discussions where care home employees have the opportunity to discuss their reasons for not getting the vaccine; this will demonstrate respect for the views of people who choose not to have a COVID-19 vaccine, while also providing an opportunity to encourage vaccination uptake. Third, consideration should be given as to the value of making COVID-19 vaccination a condition of deployment for those working in older adult care homes. Although making COVID-19 vaccination a condition of deployment may encourage some to receive the vaccination, findings from the current study suggest that, for others, making COVID-19 vaccination a condition of deployment will serve to increase vaccine hesitancy and potentially stigmatisation. Alongside the reported negative experiences of working in care during the COVID-19 pandemic, mandatory COVID-19 vaccination could result in care home employees choosing to leave their jobs or contribute to existing feelings of disenfranchisement. The need to protect vulnerable older adults in care homes must therefore be balanced carefully against any potential to lose or disenfranchise care home employees.

### Limitations

This study is based on findings from a fairly small number of interviews (*n*=10) with care home employees in the North West of England. While data saturation was reached, the small sample size should be noted and considered alongside the findings presented. Additionally, the care home employees in this study worked in the North West of England and so may not be representative of the wider UK population or care workforce, as vaccine uptake among care home employees differs regionally. Further research could explore to what extent the themes reported here are consistent across care workers in other regions, and whether themes have remained consistent as the vaccination rollout has progressed.

## CONCLUSION

Care home employees are at increased risk of contracting COVID-19, but also show lower vaccine uptake. We identified that barriers to care home employees getting the vaccine include low perceived severity of COVID-19, concerns the vaccine was not effective, concerns about the safety and speed of development of the vaccine, and mistrust in authorities. However, these barriers could be addressed by facilitating open, honest, and non-judgmental communication about the vaccine. Specifically, information provided to care home employees should include the benefits and risks associated with getting the COVID-19 vaccine, and employers should facilitate open and non-judgmental discussions where care home employees have the opportunity to discuss their reasons for not getting the vaccine. Where care home employees did identify reasons for getting the vaccine, these included wanting to protect others, severity of COVID-19, and workplace norms. Finally, all participants expressed negative attitudes towards mandatory COVID-19 vaccines, with mandatory vaccination potentially resulting in some care home employees choosing to leave their jobs and increasing negative experiences of working during the pandemic.

## Data Availability

Transcripts from the current study are available from the corresponding author on reasonable request.

## Ethics approval and consent to participate

Public Health England (PHE) Research Ethics and Governance Group (REGG) exempted this study from requiring ethical approval because the study involved interviewing care home workers in their professional capacity. No identifiable information was collected, and all participants opted into the study and provided informed consent prior to participation.

## Author contributions

All authors contributed to the design of the study. AD conducted all interviews and data analysis; CR assisted with data analysis. AD wrote the paper with CR, LJ, and HC all commented, read, and approved the final manuscript.

## Acknowledgments

We are grateful for the local authorities in the North West who aided with the recruitment for this study.

## REFERENCES

1. Ladhani SN, Jeffery-Smith A, Patel M, et al. High prevalence of SARS-CoV-2 antibodies in care homes affected by COVID-19: Prospective cohort study, England. EClinicalMedicine 2020;28:100597.

2. Ioannidis JP, Axfors C, Contopoulos-Ioannidis DG. Second versus first wave of COVID-19 deaths: shifts in age distribution and in nursing home fatalities. Environmental research 2021;195:110856.

3. Burton JK, Bayne G, Evans C, et al. Evolution and effects of COVID-19 outbreaks in care homes: a population analysis in 189 care homes in one geographical region of the UK. The Lancet Healthy Longevity 2020;1(1):e21–31.

4. Jackson LA, Anderson EJ, Rouphael NG, et al. An mRNA vaccine against SARS-CoV-2—preliminary report. New England Journal of Medicine 2020.

5. Voysey M, Clemens SA, Madhi SA, et al. Safety and efficacy of the ChAdOx1 nCoV-19 vaccine (AZD1222) against SARS-CoV-2: an interim analysis of four randomised controlled trials in Brazil, South Africa, and the UK. The Lancet 2021;397(10269):99–111.

6. Medicines and Healthcare products Regulatory Agency. 2020. UK medicines regulator gives approval for first UK COVID-19 vaccine. https://www.gov.uk/government/news/uk-medicines-regulator-gives-approval-for-first-uk-covid-19-vaccine (accessed 25 Jun 2021)

7. Kwok KO, Lai F, Wei WI, et al. Herd immunity–estimating the level required to halt the COVID-19 epidemics in affected countries. Journal of Infection 2020;80(6):e32–3.

8. Sanche S, Lin YT, Xu C, et al. High contagiousness and rapid spread of severe acute respiratory syndrome coronavirus 2. Emerging infectious diseases 2020;26(7):1470–7.

9. NHS. COVID-19 weekly announced vaccinations 29 April 2021. 2021. https://www.england.nhs.uk/statistics/wp-content/uploads/sites/2/2021/04/COVID-19-weekly-announced-vaccinations-29-April-2021-.xlsx (accessed 25 Jun 2021).

10. Tulloch JS, Lawrenson K, Gordon AL, et al. COVID-19 vaccine hesitancy in care home staff: a survey of Liverpool care homes. medRxiv [Preprint]. March 03, 2021 [cited 2021 Jun 25] https://doi.org/10.1101/2021.03.07.21252972

11. Dickerson J, Lockyer B, Moss RH, et al. COVID-19 vaccine hesitancy in an ethnically diverse community: descriptive findings from the Born in Bradford study. Wellcome Open Research 2021;6:23.

12. Murphy J, Vallières F, Bentall RP, et al. Psychological characteristics associated with COVID-19 vaccine hesitancy and resistance in Ireland and the United Kingdom. Nature communications 2021;12(1):1–5.

13. Khubchandani J, Sharma S, Price JH, et al. COVID-19 vaccination hesitancy in the United States: a rapid national assessment. Journal of Community Health 2021;46(2):270–7.

14. Schwarzinger M, Watson V, Arwidson P, et al. COVID-19 vaccine hesitancy in a representative working-age population in France: a survey experiment based on vaccine characteristics. The Lancet Public Health 2021;6(4):e210–21.

15. Soares P, Rocha JV, Moniz M, et al. Factors associated with COVID-19 vaccine hesitancy. Vaccines 2021;9(3):300.

16. Wang K, Wong EL, Ho KF, et al. Intention of nurses to accept coronavirus disease 2019 vaccination and change of intention to accept seasonal influenza vaccination during the coronavirus disease 2019 pandemic: A cross-sectional survey. Vaccine 2020;38(45):7049–56.

17. Gualano MR, Corradi A, Voglino G, et al. Healthcare Workers’(HCWs) attitudes towards mandatory influenza vaccination: A systematic review and meta-analysis. Vaccine 2021.

18. BBC. Covid vaccine to be compulsory for England care home staff. 2021. https://www.bbc.co.uk/news/uk-57492264 (accessed 25 Jun 2021).

19. Bhati KS, Hoyt WT, Huffman KL. Integration or assimilation? Locating qualitative research in psychology. Qualitative Research in Psychology 2014;11(1):98–114.

20. Carrasco JA, Lucas K. Workshop synthesis: Measuring attitudes; quantitative and qualitative methods. Transportation Research Procedia 2015;11:165–71.

21. Karafillakis E, Dinca I, Apfel F, et al. Vaccine hesitancy among healthcare workers in Europe: A qualitative study. Vaccine 2016;34(41):5013–20.

22. Goldsmith H, McCloughen A, Curtis K. The experience and understanding of pain management in recently discharged adult trauma patients: A qualitative study. Injury 2018;49(1):110–6.

23. Ritchie J, Spencer L. Qualitative data analysis for applied policy research. In: Bryman A, Burgess, RG eds. Analysing Qualitative Data. London: Routledge 1994:173–194

24. Ritchie J, Lewis J. Qualitative Research Practice. Sage Publications, London 2003.

25. Fusch PI, Ness LR. Are we there yet? Data saturation in qualitative research. The qualitative report 2015;20(9):1408.

26. Guest G, Bunce A, Johnson L. How many interviews are enough? An experiment with data saturation and variability. Field methods 2006;18(1):59–82.

27. Betsch C, Schmid P, Heinemeier D, et al. Beyond confidence: Development of a measure assessing the 5C psychological antecedents of vaccination. PloS one 2018;13(12):e0208601.

28. MacDonald NE. Vaccine hesitancy: Definition, scope and determinants. Vaccine 2015;33(34):4161–4.

29. Kwok KO, Li KK, Wei WI, et al. Influenza vaccine uptake, COVID-19 vaccination intention and vaccine hesitancy among nurses: A survey. International journal of nursing studies 2021;114:103854.

30. Dubé E, Gagnon D, Vivion M. Public Health Network: Optimizing communication material to address vaccine hesitancy. Canada Communicable Disease Report 2020;46(2-3):48.

31. Scherer LD, Shaffer VA, Patel N, Zikmund-Fisher BJ. Can the vaccine adverse event reporting system be used to increase vaccine acceptance and trust?. Vaccine 2016;34(21):2424–9.

32. Graeber D, Schmidt-Petri C, Schröder C. Attitudes on voluntary and mandatory vaccination against COVID-19: Evidence from Germany. PloS one 2021;16(5):e0248372.

33. Betsch C, Böhm R. Detrimental effects of introducing partial compulsory vaccination: experimental evidence. The European Journal of Public Health 2016;26(3):378–81.

34. de Figueiredo A, Larson HJ, Reicher SD. The potential impact of vaccine passports on inclination to accept COVID-19 vaccinations in the United Kingdom: evidence from a large cross-sectional survey and modelling study. medRxiv [Preprint]. 2021 June 1 [cited 2021 Jun 25] https://doi.org/10.1101/2021.05.31.21258122

35. Omer SB, Betsch C, Leask J. Mandate vaccination with care. 2019. https://www.nature.com/articles/d41586-019-02232-0?fbclid=IwAR0L1quO2xIBobCitNix5K-5sHa8LrTHf9NhUzVSN_CwHPiFjPh3gjmud38 (accessed 25 Jun 2021).

36. Sprengholz P, Betsch C. Herd immunity communication counters detrimental effects of selective vaccination mandates: Experimental evidence. EClinicalMedicine 2020;22:100352.

37. Osama T, Razai MS, Majeed A. Covid-19 vaccine passports: access, equity, and ethics. BMJ 2021;373:n861.

